# Instrument–service design misalignments in community age-friendly governance in China (1993–2025): A place-based analysis

**DOI:** 10.1101/2025.08.03.25332907

**Authors:** Yutian Tang, Sijing Chen

## Abstract

**Background:** Persistent mismatches between policy instruments and service design dimensions have hindered the delivery of inclusive aging services in China’s community-based governance. While aging policy agendas increasingly reference integration and user-centred care, the operational tools remain biased toward infrastructural and regulatory interventions. This study addresses the gap by linking policy instrument theory with design-informed service logic.

**Methods:** A two-dimensional analytical framework was developed, mapping 22 policy instruments to six service design dimensions—procedural integration, emotional support, participatory engagement, informational communication, institutional coordination, and strategic goal setting. Based on 48 national-level aging policy documents issued from 1993 to 2025, we conducted NVivo-assisted coding, word frequency mapping, and co-occurrence heatmap analysis. Fieldwork in Zhenzhu Community (Feixi County) further substantiated institutional coordination patterns and local service implementation dynamics. This framework was applied to identify and analyze coordination issues through empirical data from policy documents and non-participant observation.

**Results:** The analysis identified a path-dependent reliance on supply-driven and environmental instruments, with limited attention to demand-responsive and affective elements. Three key coordination failures were identified: procedural fragmentation, emotional neglect, and cross-sectoral misalignment. Empirical findings from Zhenzhu Community highlighted how these mismatches weaken service continuity, marginalise older adults’ lived experiences, and restrict adaptive planning.

**Conclusion:** By integrating service design principles into instrument analysis, the study reframes how institutional configurations shape aging governance. A recalibrated policy instrument mix—one that embeds feedback loops, promotes participatory mechanisms, and strengthens interdepartmental collaboration—is essential for enabling adaptive and user-centred service systems. The proposed framework offers transferable insights for policy reforms in other lower- and middle-income countries facing parallel demographic and governance challenges.

## 1. Introduction

Population aging has emerged as a defining demographic and governance challenge worldwide. By 2050, one in six individuals globally is projected to be aged 65 or above[1]. In China, this demographic transformation is particularly pronounced: by the end of 2024, the population aged 60 and above had reached 310 million (22% of the total), including 220 million aged 65 and over (15.6%)[2]. In response, a series of national policy initiatives—such as the Healthy China 2030 Planning Outline[3] and the 14th Five-Year Plan for the Development of National aging Undertakings and the Elderly Care Service System[4]—have aimed to improve the inclusiveness, adaptability, and spatial responsiveness of community-based aging services. However, the policy response has remained primarily infrastructure-led and fragmented, with limited attention to how specific institutional instruments shape service experiences at the emotional, participatory, and spatial levels. Research has shown that neighbourhood environments, including green space availability and socio-economic accessibility, significantly affect older adults’ cognitive and emotional well-being[5]. Nevertheless, such spatial determinants are rarely embedded into Chinese aging policies as actionable governance mechanisms.

Service design offers a framework for translating aging policies into practice through spatial, emotional, and participatory elements[6]. In addition, synthesising insights from design studies and aging governance, this study operationalises six interrelated service design dimensions: spatial accessibility, procedural integration, emotional support, participatory engagement, informational communication, and institutional coordination[7][10]. These dimensions move beyond standardised infrastructure provision by foregrounding user-centred, affective, and co-productive aspects of service delivery. Although growing global attention has been paid to place-sensitive and person-centred approaches[11]-[12], China’s aging governance remains constrained by a path-dependent reliance on infrastructural investments and regulatory mandates.

This study adopts a policy instrument perspective to examine how the design and deployment of instruments condition the delivery of aging services. In particular, drawing on Salamon’s conceptualisation of policy tools as institutionalised mechanisms for resource allocation and behavioural regulation, policy instruments are viewed not only as functional levers but also as normative devices that encode policy priorities into practice[13]. Moreover, the classification by Rothwell and Zegveld—categorising instruments as supply-, demand-, and environment-oriented—is adapted here to reflect the layered and sectoral nature of China’s community governance system[14]. When applied to the context of aging governance, this typology enables the identification of tool-path dependencies and their implications for service design.

To bridge the theoretical divide between policy implementation and service experience, this study establishes a two-dimensional analytical framework linking 22 types of policy instruments to six service design dimensions. The framework is grounded in NVivo-assisted qualitative content analysis of 48 national-level policy documents issued between 1993 and 2024[15]-[17]. A co-occurrence heatmap and fieldwork findings from Zhenzhu Community in Hefei, Anhui Province, are used to empirically validate patterns of tool usage and their implications for service delivery.

Globally, aging policies in countries such as Japan, Singapore, and the United Kingdom have increasingly incorporated behavioural insights, participatory feedback loops, and design-based frameworks to improve service accessibility and intersectoral coordination[18][19]. These international shifts suggest that service design thinking and tool-based governance are not mutually exclusive, but rather synergistic when aligned. However, in China, the limited integration of participatory and emotional mechanisms remains a structural constraint that marginalises lived experiences of older adults. Against this backdrop, the present study makes two core contributions. First, it offers a design-informed framework for re-evaluating aging policy. Second, it empirically identifies coordination gaps and emotional blind spots within China’s current policy landscape. By tracing the misalignments between tools and service design dimensions, this study contributes to international scholarship on aging, design, and governance, while offering actionable insights for policy optimisation in similarly situated contexts[20].

## 2. Materials and Methods

### 2.1 Policy Samples and Data Sources

This study examined the alignment between policy instruments and service design dimensions by compiling a dataset of national-level policy documents issued between 1993 and 2025. The starting year of 1993 was selected because it marked the issuance of China’s first national policy that explicitly emphasised both “community” and “service provision” at the central government level—namely, the Opinions of the State Council on Accelerating the Development of Community Service Industries[21]. This document, in particular, signalled the institutional emergence of aging-oriented service delivery within China’s policy framework. A total of 187 documents were initially retrieved through keyword-based searches on official platforms, including those of the State Council, the Ministry of Civil Affairs, and the National Health Commission. Keywords such as “aging,” “community services,” “elderly care,” “public service delivery,” and “urban renovation” were employed to ensure thematic relevance to both aging governance and Service Design concerns[22]. A three-step screening procedure was adopted to ensure analytical rigour and thematic consistency. First, only documents formally issued or jointly endorsed by the State Council or its directly affiliated ministries were retained, ensuring institutional authority. Second, documents were retained only if they contained explicit references to community-based aging services, such as infrastructure adaptation, health–social integration, service delivery governance, or spatial planning strategies. Third, documents solely concerned with institutional eldercare, rural development, or general welfare policy—without an aging-specific orientation—were excluded. This selection process yielded a final sample of 48 national-level policies for qualitative content analysis.

### 2.2 Fieldwork in Zhenzhu Community

To contextualise the national policy analysis with grounded empirical evidence, fieldwork was conducted in Zhenzhu Community, a peri-urban neighbourhood in Feixi County, Hefei City, Anhui Province, China. The community has a registered population of 9,041, of whom 1,194 individuals (13.2%) are aged 60 and above[23]. As a rapidly urbanising area at the intersection of spatial governance and demographic transformation, Zhenzhu Community reflects common challenges faced by aging populations in transitional urban settings.

Unstructured Interviews were carried out with local community staff to examine practical aspects of service coordination, infrastructure adaptation, and older adult engagement. The aim was to explore how national aging-related policies are interpreted and operationalised at the local level. No personal or sensitive data were collected from residents during fieldwork. In line with institutional guidelines, the research was deemed exempt from formal ethics review procedures.

### 2.3 Identification and Classification of Policy Instruments

An inductive–deductive coding strategy, guided by Rothwell and Zegveld’s typology, facilitated the identification of 22 distinct policy instruments. These were categorised into three governance orientations: supply-oriented, environmental-oriented, and demand-oriented. The classification process emphasised conceptual clarity and operational specificity, ensuring that each instrument demonstrated cross-document recurrence and functional relevance. Coding was independently conducted by two researchers and validated through iterative consensus.

The resulting typology constitutes the horizontal axis of the analytical framework, with detailed definitions presented in Table 1.

**Table 1.** Classification of 22 policy instruments by functional orientation and their implications for community aging services.

| Type | Instrument Name | Functional Implication in Community aging Services |
| --- | --- | --- |
| Supply-oriented | Organisational Leadership | Coordinates cross-sectoral collaboration and ensures institutional accountability in aging service delivery. |
|  | Public Services Provision | Enhances accessibility and efficiency of facilities tailored to older adults’ daily needs. |
|  | Infrastructure Development | Upgrades physical environments to meet mobility, housing, and care demands of the elderly. |
|  | Digital Infrastructure | Promotes digital platforms with age-friendly interfaces to improve service navigation and delivery. |
|  | Workforce Development | Trains professionals across healthcare, rehabilitation, and ICT to strengthen service continuity and quality. |
|  | Capital Investment | Channels fiscal inputs (e.g., lottery welfare funds) to support infrastructure renovation and service |
|  |  | expansion. |
| Environmental<br>-oriented | Legal and<br>Regulatory<br>Supervision | Establishes regulatory frameworks to safeguard the rights and dignity of older adults. |
|  | Service<br>Standards and<br>Norms | Defines benchmarks for quality assurance, safety, and equity in community-based elderly services. |
|  | Value<br>Orientation | Cultivates a culture of respect, intergenerational harmony, and positive aging through moral and civic education. |
|  | Financial<br>Support | Facilitates elderly access to inclusive financial mechanisms, insurance coverage, and credit access for care-related expenses.. |
|  | Strategic Goal<br>Setting | Articulates policy objectives and long-term visions for age-friendly community transformation. |
|  | Rights and<br>Interests<br>Protection | Provides subsidies and enforcement mechanisms to uphold welfare entitlements for seniors. |
|  | Pilot Programs<br>and<br>Demonstration<br>Projects | Tests innovative service models and scales up replicable community practices. |
|  | Age-Friendly<br>Renovations | Implements barrier-free retrofits to improve residential safety and environmental usability. |
|  | Publicity and<br>Education | Raises public awareness and civic participation in aging service initiatives. |
| Demand-oriented | Industrial<br>Upgrading and<br>Product<br>Research and<br>Development | Stimulates innovation of aging-adapted products and market-oriented service ecosystems. |
|  | Social<br>Participation | Encourages diverse actors (e.g., NGOs, volunteers) in co-producing aging services and fostering community ties. |
|  | Social Welfare<br>Provision | Strengthens the subsidy system to ensure fulfilment of basic needs and reduce service exclusion for vulnerable older adults. |
|  | Elder Support<br>and Income<br>Protection | Enhances financial protections and allowances to support independent living and health security. |
|  | Healthcare<br>Security | Integrates healthcare with elderly care to ensure continuity of medical and preventive services. |
|  | Procurement<br>and Service<br>Outsourcing | Delegates service provision to qualified third parties<br>to improve delivery efficiency and responsiveness. |
|  | Needs<br>Assessment | Identifies differentiated user needs to guide targeted<br>service design and resource allocation. |

This table categorises instruments into three functional types—supply-oriented, environmental-oriented, and demand-oriented—and summarises their respective roles in facilitating aging service delivery at the community level. The classification is based on inductive coding of national policy documents and adapted to reflect service design relevance.

### 2.4 Analytical Framework and Coding Structure

Guided by Rothwell and Zegveld’s typology, this study defines the horizontal axis of the framework as comprising 22 distinct policy instruments. These are categorised into three governance orientations: supply-oriented instruments (e.g. infrastructure, funding, human resources), environmental instruments (e.g. regulations, standards, strategic planning), and demand-oriented instruments (e.g. participation, needs assessment, service customisation). The vertical axis was developed through inductive thematic synthesis of aging-related policy texts, informed by established service design principles[24]. In particular, six core service design dimensions were identified: spatial accessibility, procedural integration, emotional support, participatory engagement, informational communication, and institutional coordination. These six dimensions, in fact, are grounded in established service design literature[25]. They are based on human-centred and co-creative principles, and were adapted in this study to fit the context of community-based aging governance in peri-urban China. The spatial accessibility dimension, for example, was defined as the extent to which age-friendly policies incorporate physical environment design, including accessibility, mobility, and neighbourhood features. This dimension, moreover, draws on prior research linking neighbourhood environments with health and active aging outcomes[26].

Coding reliability was ensured through independent double-coding and consensus validation, following established recommendations for improving intercoder agreement[27]. This coding structure underpins the co-occurrence heatmap introduced in Section 3.2, offering a visual representation of alignment patterns between instruments and design dimensions.

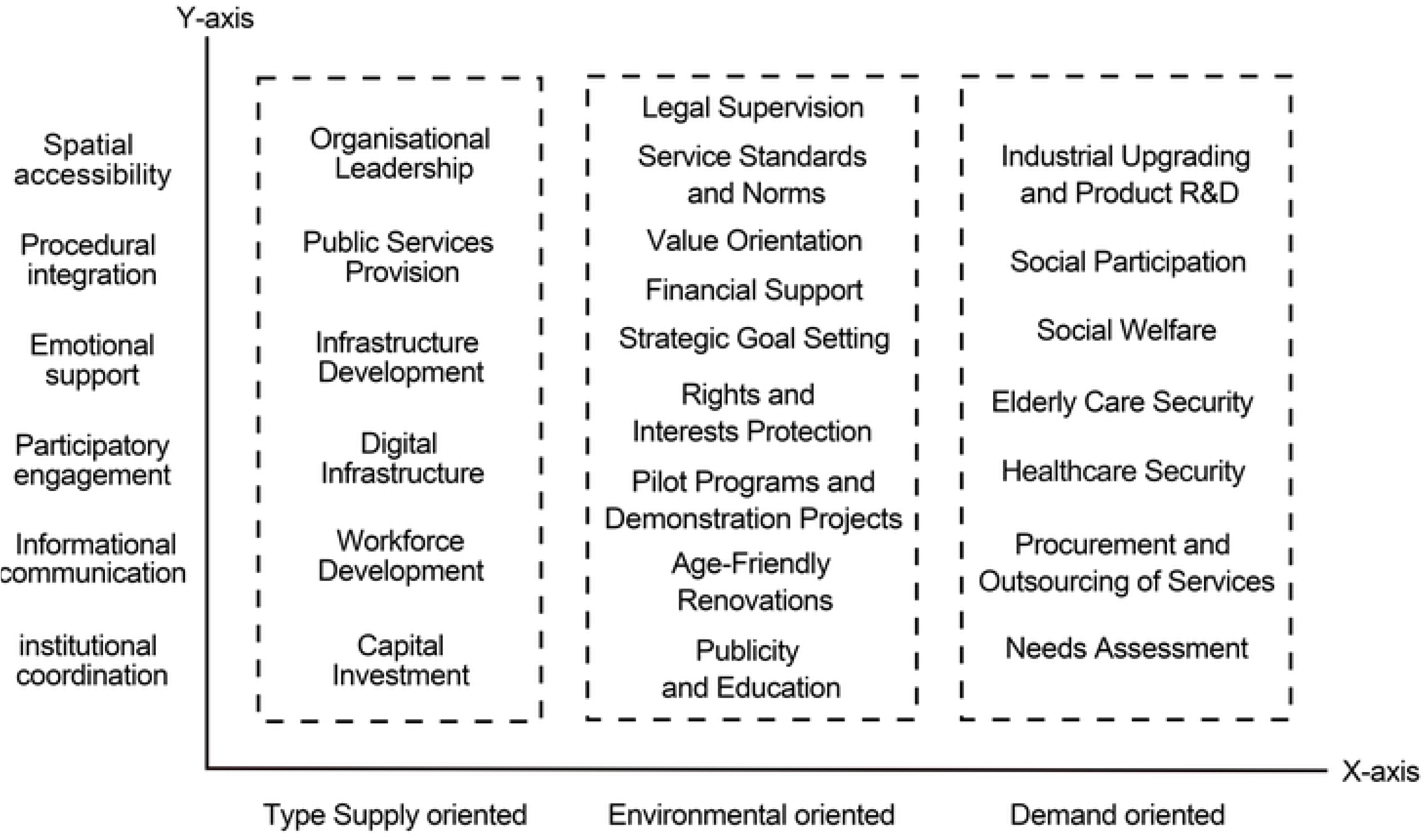

Fig 1 illustrates the full analytical matrix, where 22 policy instruments (horizontal axis) intersect with six service design dimensions (vertical axis). Each cell represents a co-occurrence point, allowing systematic assessment of alignment or mismatch between policy tools and service delivery elements.

### 2.5 Instrument–Design Alignment Mechanisms

To clarify the operational relationships between policy instruments and service design dimensions, this section introduces the theoretical coordination mechanisms that underpin the analytical framework. Rather than treating tools and dimensions as independent variables, the analysis conceptualizes them as interactive components in the governance of aging services. This study explores the interaction between different categories of policy instruments—supply-oriented, environmental-oriented, and demand-oriented—and service design dimensions, examining how these interactions shape the effectiveness of community-based aging governance. Because these instruments influence the delivery of specific services, they can either enable or constrain the overall service design. Therefore, the analysis investigates the ways in which these instruments affect service functions and their interaction with design dimensions. This coordination occurs through three main pathways: instrumental activation, feedback modulation, and institutional integration. Building on Salamon’s theory of policy tools, these instruments are conceptualized as institutional devices for shaping behavior and redistributing resources[13].

Supply-oriented instruments primarily activate structural service functions. These tools support dimensions like spatial accessibility and procedural integration by creating the physical and administrative conditions necessary for service provision. For example, investment in age-friendly facilities enables spatial access, while staffing guidelines influence the routinization of service procedures.

Environmental-oriented instruments shape the normative and legal environment in which services operate. These tools promote institutional integration and procedural alignment but may constrain emotional responsiveness or participatory engagement. Their top-down design can institutionalize consistency but often lacks sensitivity to lived experience or individualized needs.

Demand-oriented instruments enhance feedback channels and emotional alignment. These tools engage older adults directly in shaping service content and delivery processes. When implemented effectively, they strengthen emotional support, informational communication, and procedural flexibility.

Each tool-dimension interaction was classified based on its functional role: enabling, shaping, or restricting a given design function. Table 2 visualizes this coordination matrix, offering a conceptual map of how instruments influence service quality and responsiveness. This mapping process forms the theoretical basis for the co-occurrence analysis presented in Section 3.

**Table 2.**
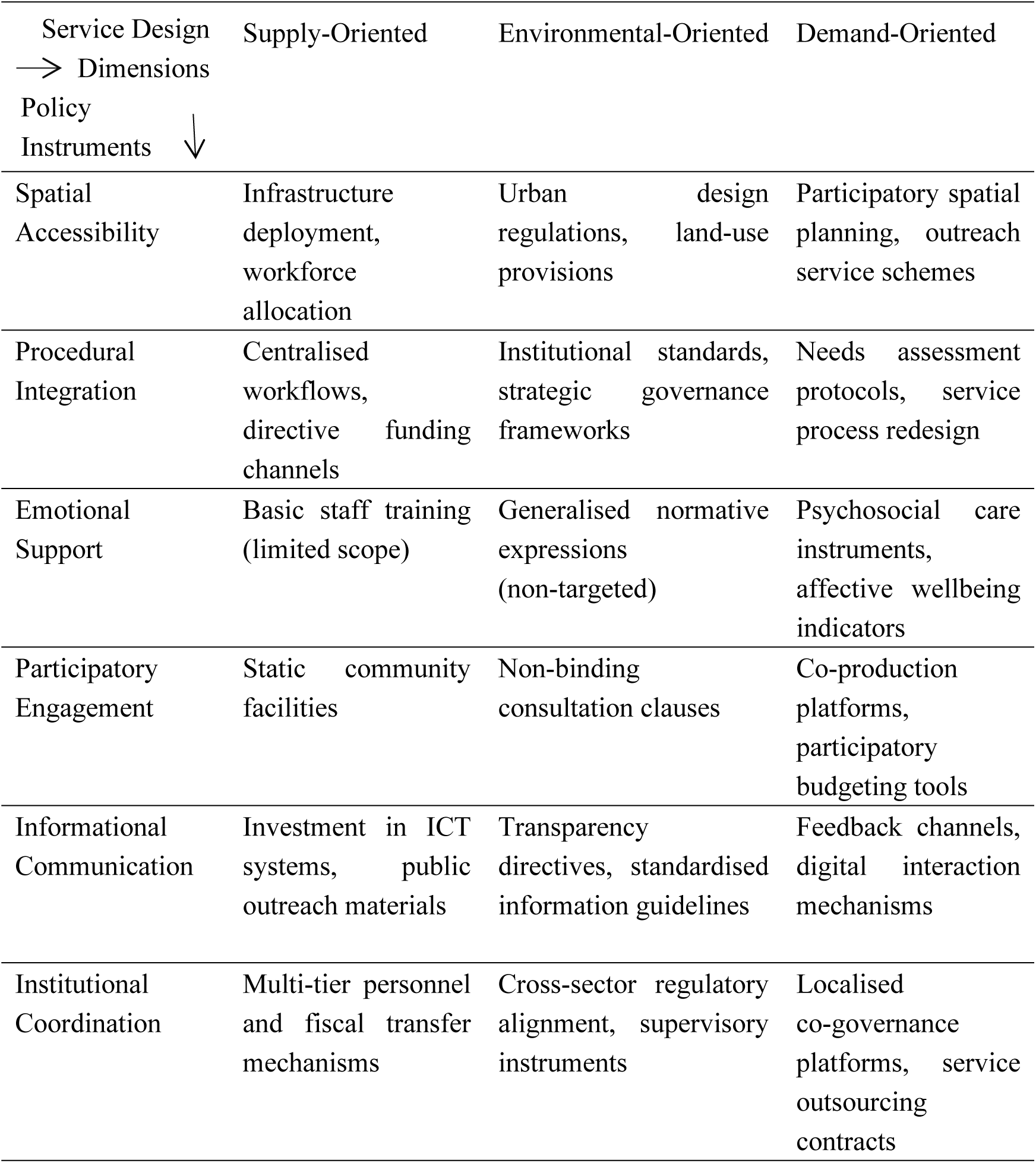
Theoretical Mapping of Policy Instruments to Service Design Dimensions.

This table presents the hypothesised coordination pathways between three categories of policy instruments (supply-oriented, environmental-oriented, and demand-oriented) and six core service design dimensions. It serves as a conceptual foundation for the coding schema and co-occurrence analysis introduced in later sections.

## 3. Results

### 3.1 Structural Imbalance in the Deployment of Policy Instruments

Analysis of the 48 national-level policy documents reveals a clear structural imbalance in the deployment of policy instruments within age-friendly community governance. Supply-oriented instruments constituted 37.97% of the coded references, while environmental tools accounted for the largest share at 41.41%. In contrast, demand-oriented instruments represented only 20.62% of all coded content. This asymmetry is detailed in Table 3, which summarises the frequency distribution of the 22 identified policy tools across the three functional categories

**Table 3.**
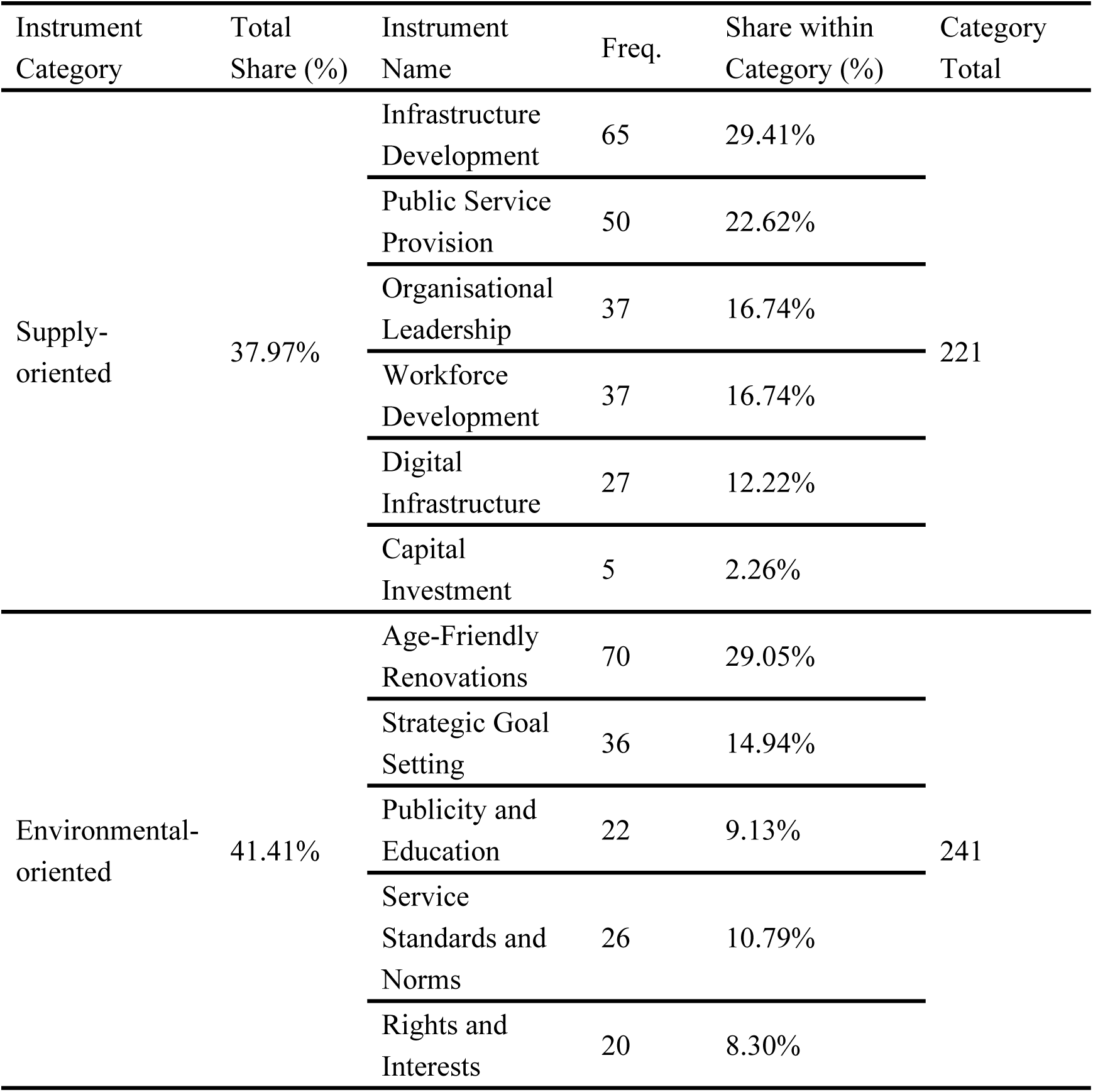
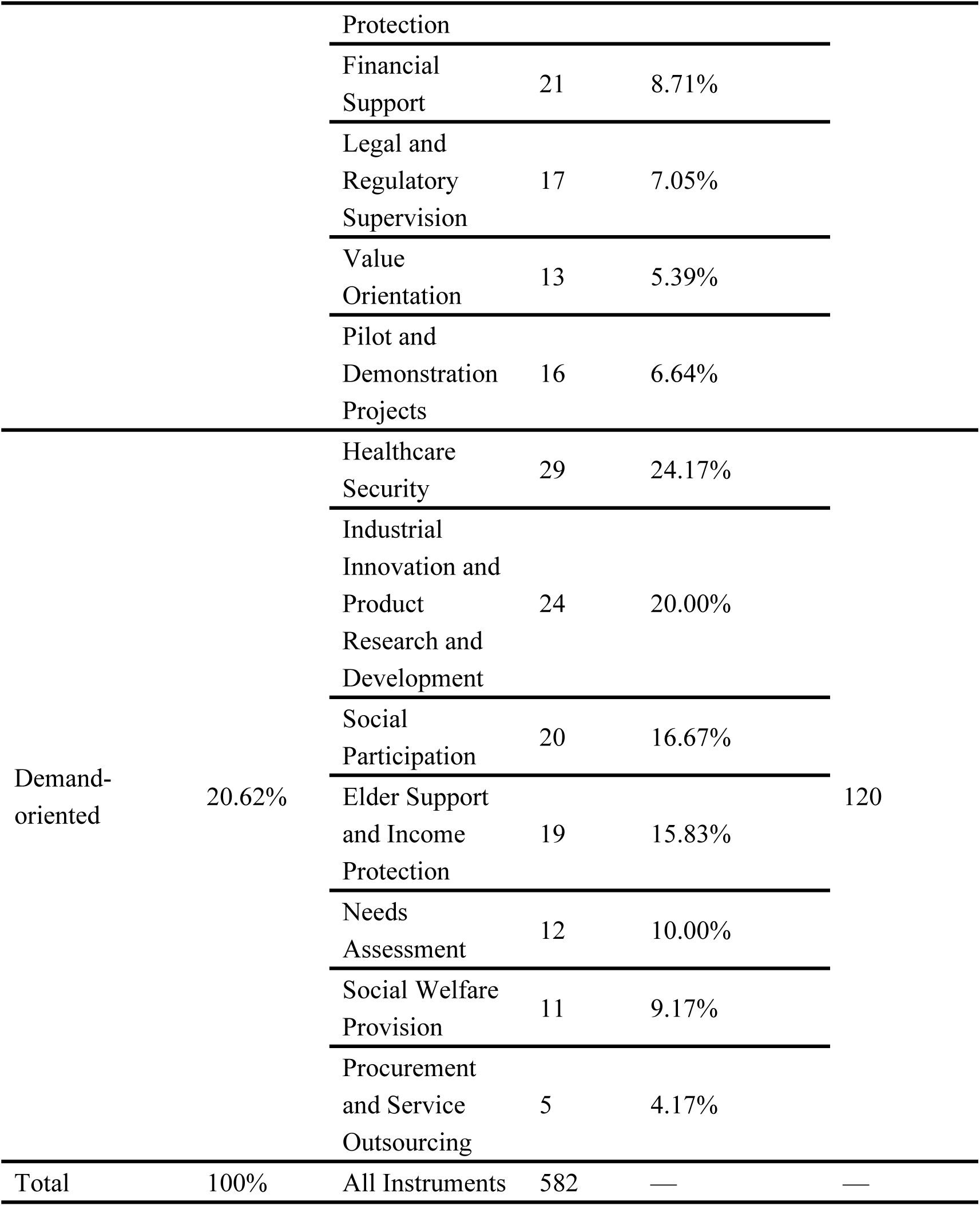
Frequency Distribution of 22 Policy Instruments by Functional Category.

This table presents the frequency and proportional share of 22 identified policy instruments across three functional categories—supply-oriented, environmental-oriented, and demand-oriented—based on NVivo coding of 48 national-level policy documents. “Freq.” indicates the total number of coding references per instrument. “Category Total” refers to the sum of frequencies within each functional category.

This distribution reflects an entrenched governance logic that privileges infrastructure-led expansion and top-down administrative control. Supply-oriented instruments enable rapid facility development and centralised resource mobilisation, yet they lack sensitivity to local variation and user heterogeneity. Environmental instruments support regulatory consistency and strategic alignment, but often fall short in stimulating behavioural shifts or fostering co-productive engagement at the community level.

By contrast, demand-oriented instruments—which underpin responsiveness, emotional well-being, and participatory inclusion—remain institutionally marginalised. Tools such as “Needs Assessment,” “Social Welfare Provision,” and “Procurement and Service Outsourcing” were mentioned infrequently, indicating a critical gap in instruments aimed at enabling market participation and empowering local actors.

This structural imbalance not only constrains the adaptability of China’s aging governance framework but also undermines the foundation for place-based interventions that address spatial and emotional inequalities in later-life health. Previous research has highlighted that advancing relational aging requires not only spatial adaptation but also institutional commitment to emotional well-being and participatory service environments, as aging-in-place involves the co-construction of affective and temporalised experiences of place. These findings underscore the need to recalibrate the current policy instrument mix toward more adaptive, inclusive, and emotionally responsive models of aging service delivery.

### 3.2 Misalignment Between Policy Instruments and Service Design Dimensions

#### 3.2.1 NVivo-Based Matrix Analysis

As shown in Fig. 2, the heatmap visualises the co-occurrence frequencies between 22 policy instruments and six service design dimensions, derived from NVivo matrix coding of 48 national policy documents. In particular, the heatmap highlights the frequency with which certain tools intersect with specific dimensions, providing a clear visual representation of the alignment between policy tools and service functions. The darker cells, in turn, indicate stronger interactions between tools and dimensions, while lighter cells represent weaker or less frequent interactions. This visual representation, moreover, facilitates the identification of key alignment or misalignment patterns, which are critical for understanding the effectiveness of current policy designs in addressing service needs.

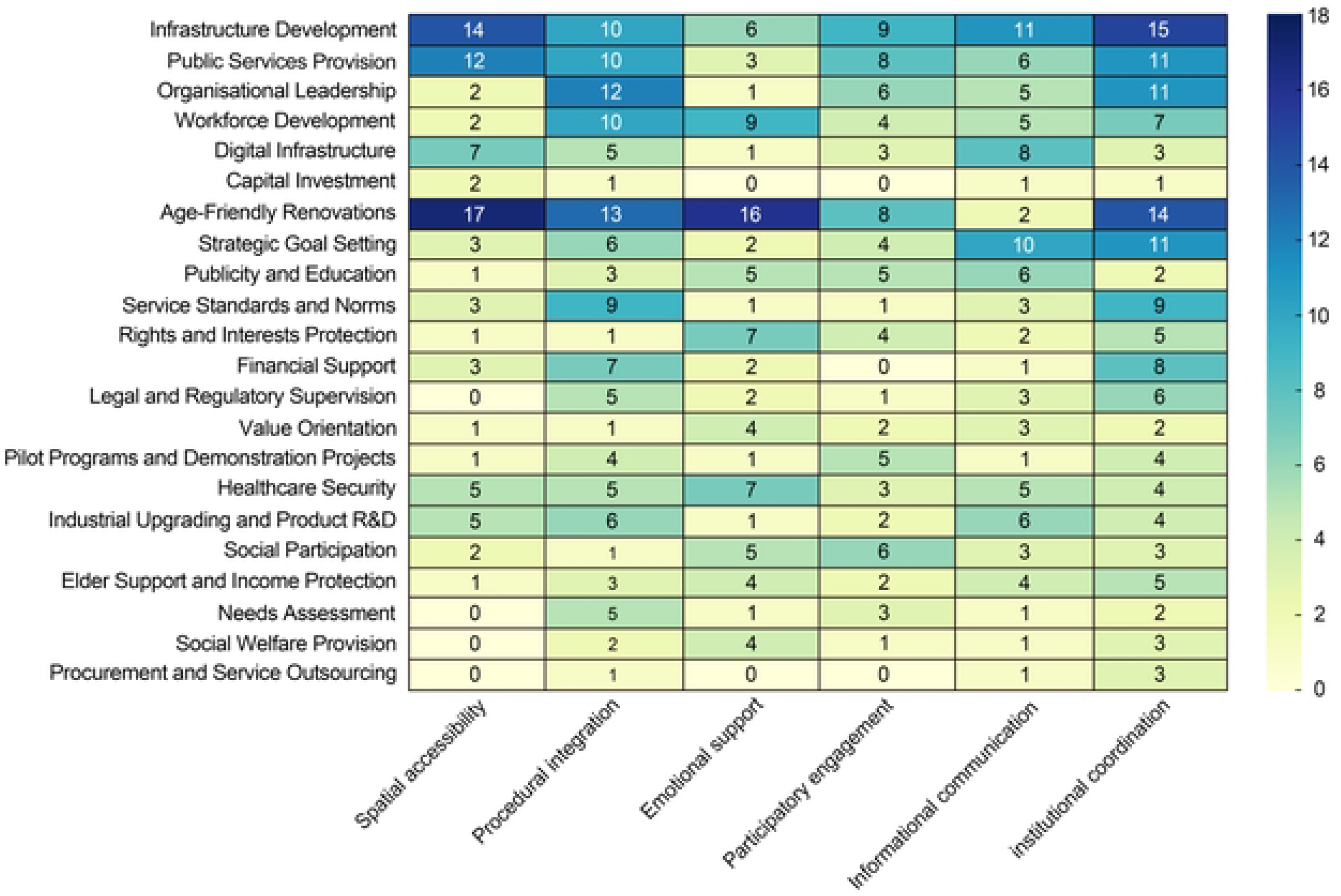

A clear pattern of structural misalignment emerges from the distribution. Supply-oriented instruments—such as Infrastructure Development (n=14 in spatial accessibility; n=15 in institutional coordination), Public Service Provision (n=12 and n=11)—exhibit strong linkages to procedural or infrastructural domains. However, their intersections with emotionally responsive and participatory elements—such as emotional support or participatory engagement—remain significantly weaker.

Environmental tools, particularly Age-Friendly Renovations (n=16 in emotional support), present relatively better alignment with emotional responsiveness. Yet tools such as Strategic Goal Setting and Publicity and Education seldom appear alongside participatory mechanisms or user feedback loops. Demand-oriented instruments—including Needs Assessment, Social Welfare Provision, and Procurement and Outsourcing—show sparse connections across all six dimensions, with frequencies ranging from 0 to 5.

This visual structure reinforces a systemic imbalance: policy implementation still disproportionately favours institutional visibility and physical infrastructure, rather than relational care, iterative design, or co-productive service innovation. The marginal alignment with dimensions such as emotional support, participatory engagement, and informational communication points to a persistent neglect of affective and user-informed aspects of aging service systems.

#### 3.2.2 Field-Based Observations from Zhenzhu Community

Field-based observations were conducted in January 2025 in Zhenzhu Community, involving semi-structured interviews with service providers. Empirical insights from fieldwork in Zhenzhu Community further corroborate the misalignment revealed by the policy corpus. Although staff reported improvements in spatial accessibility—such as ramps, widened pathways, and accessible signage—there remained significant gaps in emotionally attuned care, community responsiveness, and participatory governance.

Observations revealed limited continuity in older adult engagement activities, weak interdepartmental linkages, and an absence of digital or psychosocial support adapted to the diverse needs of elderly residents. While information campaigns and planning targets were frequently observed during field visits, these initiatives rarely involved iterative user feedback or cross-sectoral coordination. In addition, policy discourse frequently references terms like “mental well-being” and “intergenerational harmony,” yet these remain largely rhetorical, with few mechanisms for budget allocation, performance evaluation, or integrated digital platforms. This illustrates what Béland terms the rhetorical–instrumental gap, where aspirational language fails to generate operational change[28].

#### 3.2.3 Keyword Frequencies and Policy Discourse

Supporting this observation, Table 4 summarises the ten most frequently occurring keywords across all documents. These are largely institutional or service-driven terms (e.g., Service, Community, Elder Care), while emotionally grounded or participatory expressions (e.g., empathy, co-design, feedback) are notably absent. This pattern reinforces the marginalisation of user-centred perspectives in aging governance.

**Table 4.** Top 10 Keywords in aging-Related Policy Documents (1993–2025)

| No. | Keywords | Weighted Proportion of Keywords |
| --- | --- | --- |
| 1 | Service | 4.33 |
| 2 | Elder Care | 4.14 |
| 3 | Older Adults | 2.04 |
| 4 | Community | 4.05 |
| 5 | Society | 0.96 |
| 6 | Institution | 0.83 |
| 7 | Age | 0.74 |
| 8 | Standard | 0.72 |
| 9 | Facilities | 0.68 |
| 10 | Construction | 0.60 |

This table lists the ten most frequently referenced keywords extracted from 48 national-level policy documents, based on NVivo word frequency analysis. The “Weighted Proportion of Keywords” reflects the relative frequency of each term after normalisation by document length. Keywords such as “Service,” “Community,” and “Elder Care” dominate, whereas emotionally expressive or co-creative terms are notably absent.

Taken together, the heatmap, field interviews, and keyword analysis converge to reveal a systematic bias in instrument deployment. This misalignment weakens the integration of human-centred service principles into aging policy and hampers the development of place-based, adaptive, and affective service ecosystems essential for later-life well-being. This instrumental gap hinders the development of health-responsive, spatially adaptive aging services.

### 3.3 Temporal Evolution in Policy Orientation

To further explore the systemic misalignment between policy instruments and service design dimensions, it is critical to examine the evolution of governance orientations over time. In particular, drawing upon 48 national policy documents, this study divides China’s age-friendly governance trajectory into three distinct phases: Initial Development Phase (1993–2010), Rapid Growth Phase (2011–2020), and Optimization and Upgrading Phase (2021–Present). These stages reflect the gradual transformation in policy focus, the increasing complexity of governance tools, and the evolving integration of service design principles into ageing policies.

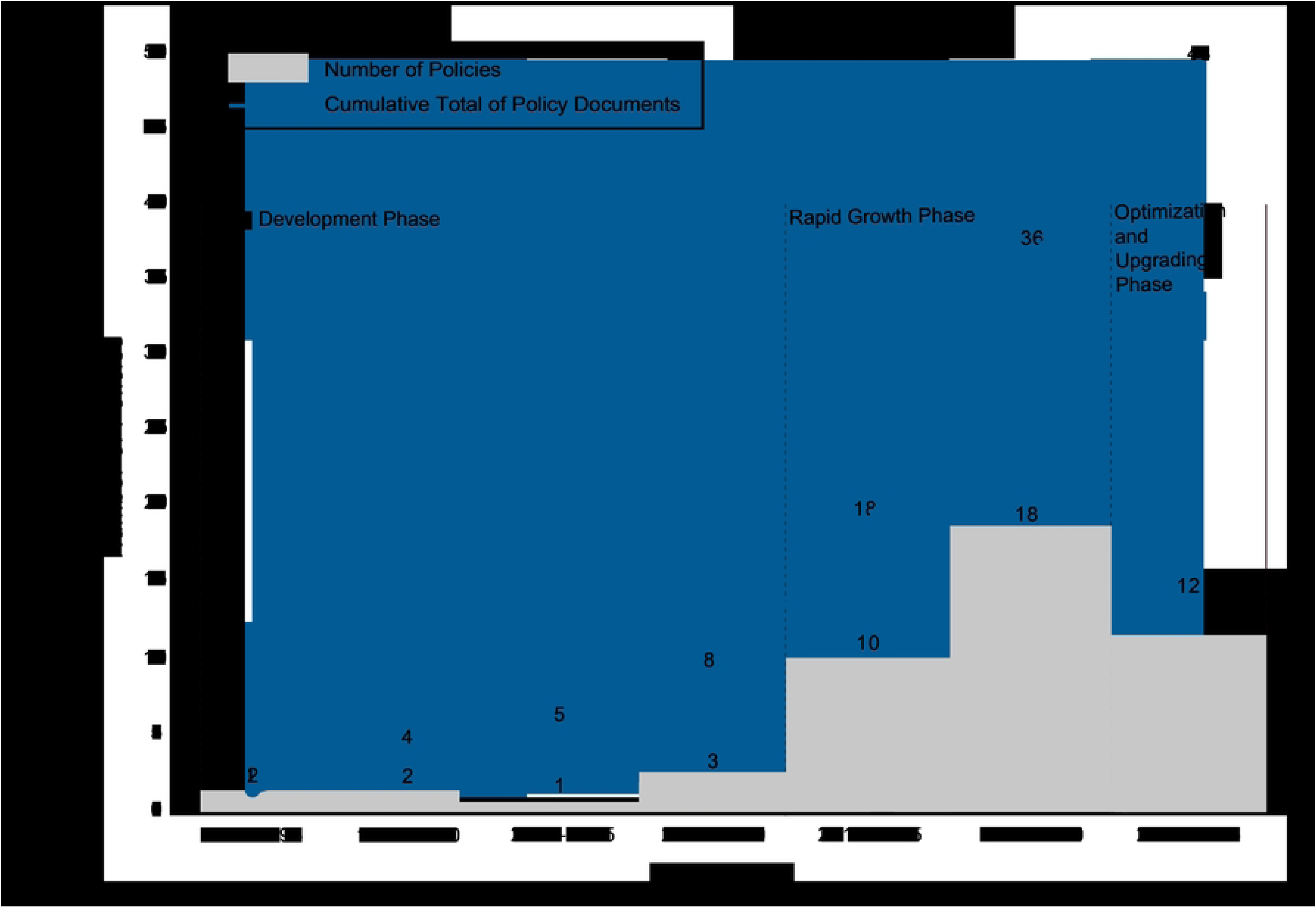

Temporal distribution and cumulative count of national ageing-related policy documents (1993–2025), segmented into three governance phases, illustrating shifts in instrument type and the ongoing integration of service design principles.

During initial Development Phase, China’s ageing policy framework began to take form, although its engagement with ageing issues remained limited. Policy instruments primarily operated within the broader context of welfare agendas, often lacking a dedicated focus on service design elements. The 2000 State Council guideline introduced the principle of “home-based, community-supported, institution-complemented” care, yet it failed to provide concrete tools or participatory mechanisms to support this shift[29]. As a result, the phase is characterized by a symbolic commitment to ageing governance without substantial operational mechanisms, leaving family-based care as the predominant model while community and institutional services played a supplementary role. The emergence of these foundational policies represents a recognition of ageing challenges, but the tools employed during this period were insufficient to catalyze significant structural change.

Rapid Growth Phase marked a surge in policy activity, characterized by a notable increase in the number of ageing-related policy documents. The year 2011 saw the issuance of the “12th Five-Year Plan for the Development of Ageing Undertakings,” which proposed an integrated system based on home care, supported by community services and bolstered by institutional care[30]. Rhetorical shifts emphasized “community empowerment” and “integrated service delivery,” suggesting an ideological transformation in the approach to elderly care.

However, despite the broadening scope of policies, the actual implementation remained heavily reliant on supply-oriented and environmental tools, with insufficient attention given to demand-driven and participatory mechanisms. Although community engagement was increasingly referenced, the development of tools for participatory design, informational communication, and institutional coordination remained underdeveloped, limiting the practical impact of the policy shift. This phase illustrates the rapid expansion of policy activity, but with a persistent reliance on traditional, infrastructure-focused governance tools. The ideological shift towards integrated services was not yet matched by the institutionalization of participatory and user-centered practices.

In optimization and Upgrading Phase, China’s ageing policies have increasingly focused on enhancing the quality and inclusiveness of services, moving beyond mere service expansion. The “14th Five-Year Plan” (2021) introduced a focus on the development of home-based care beds and on-demand services[31]. The integration of “Big Health” concepts, along with advancements in information technology and age-friendly innovations, has helped shape a more comprehensive and high-quality elderly care system.

While policy language has increasingly referenced emotional support, digital inclusion, and co-creation mechanisms, institutional mechanisms like budget allocations and coordination platforms have struggled to keep pace. These gaps reflect what Béland terms a “rhetorical– instrumental gap,” where progressive language does not necessarily alter entrenched governance routines. This phase is marked by the slow but steady maturation of the policy landscape, emphasizing the optimization and upgrading of the existing infrastructure. Despite the growing recognition of emotional, participatory, and health-responsive services, these areas still remain peripheral in policy implementation.

The evolution of ageing policies in China reflects a gradual layering of priorities, rather than a dramatic paradigm shift. From the infrastructure-led logic that dominated the Initial Development and Rapid Growth phases, policies are now shifting toward a more inclusive and quality-focused approach in the Optimization and Upgrading phase. Yet, emotional, participatory, and health-responsive services remain underdeveloped, continuing to constrain the overall effectiveness of the ageing service system. This evolving trajectory highlights the need for a deeper, more systematic integration of service design principles to ensure that ageing policies address the full spectrum of older adults’ needs.

## 4. Discussion

### 4.1 Institutional Logics Behind Instrumental Imbalance

The heatmap and frequency matrix presented in Sections 3.1 and 3.2 reveal a clear dominance of supply-oriented instruments in China’s community-level aging policy. These instruments primarily target the provision of infrastructure, emphasise regulatory enforcement, and promote standardised procedures. This pattern reflects a consistent preference for tools that deliver measurable and visible outputs, predominantly supporting spatial accessibility and administrative efficiency. However, other service design dimensions—such as emotional support, participatory engagement, and personalisation—remain structurally marginalised. As noted by Urbaniak and Walsh, theoretical models integrating place, social exclusion, and critical life transitions remain underdeveloped in aging research[32]. This gap reinforces the urgency for policy systems to embed design-literate instruments that explicitly address spatial and social dislocation.

Even in progressive welfare regimes such as Norway, aging-in-place strategies are often framed through idealised notions of home and familial care, overlooking emotional and relational complexities embedded in later-life governance[33]. Longitudinal evidence from New Zealand further suggests that residential mobility—especially when compounded by area-level deprivation—reinforces mental health disparities through bi-directional mechanisms[34]. These international findings echo our analysis of Chinese national policies, where the operationalisation of aging support similarly neglects affective infrastructures and assumes household-based care provision.

This configuration reflects a deeply rooted institutional logic that prioritises visibility and auditability over experiential quality[35]. Echoing the “targets approach” articulated by Hood in his critique of new public management[36], Chinese policy tends to equate success with measurable outputs, reinforcing top-down hierarchies. As a result, the inclusion of aging-friendly rhetoric in policy texts often fails to translate into operational instruments capable of addressing psychosocial vulnerability or contextual heterogeneity[37].

The systematic underuse of demand-side instruments—such as participatory planning, emotional support systems, and user-informed evaluation—also points to a broader inertia in adaptive governance. As a result, without institutionalised feedback mechanisms, aging services tend to become overly standardised and technocratically driven, therefore reducing their sensitivity to the lived experience of older adults.

### 4.2 Community Evidence: Spatial Gains, Relational Gaps

Empirical fieldwork in Zhenzhu Community, Feixi County, Anhui Province, substantiates this imbalance. Although 13.2% of its population is aged 60 or above, the most visible policy outcomes are physical improvements—such as accessible pavements and barrier-free facilities—enabled by centrally deployed infrastructure programmes.

However, long-term field observations and records reveal persistent deficiencies in emotional and participatory services. Initiatives like health talks and social gatherings are often short-term, lacking continuity, feedback loops, or dedicated coordination. Crucially, no regular mechanisms exist to incorporate older residents’ voices into service design or evaluation, revealing an absence of participatory infrastructure.

These findings highlight a spatial–relational disconnect. Physical enhancements have not been accompanied by emotional support or inclusive governance, creating an environment that is superficially age-friendly but structurally fragmented. Zhenzhu thus exemplifies the broader tendency for policy tools to favour visibility over relational value.

To address these relational deficits, Zhenzhu Community could incorporate low-cost, demand-side instruments that embed emotional and participatory functions into existing service structures. Tools such as co-design workshops, elder feedback panels, and peer support groups may be implemented through regular administrative routines and modest resource allocations. Although less visible than infrastructure investments, such tools yield high relational value, directly addressing service design dimensions currently overlooked in China’s policy mix. Their institutionalisation would not only strengthen emotional resilience and social cohesion, but also help reframe aging services as participatory, dignity-centred processes—thus contributing to the correction of China’s broader instrumental imbalance.

### 4.3 International Insights and Policy Lessons: Embedding Emotional and Participatory Instruments

Comparative evidence from Japan, Singapore, and the United Kingdom illustrates alternative pathways that embed emotional infrastructure and design-literacy into policy instruments.

In Japan, the Community-Based Integrated Care System enables decentralised coordination of medical, emotional, and social services at the municipal level. Its success lies in multi-layered governance and neighbourhood-scale responsiveness, particularly in dementia care and long-term support[38].

Singapore’s Active aging Hubs integrate digital learning, psychosocial services, and community healthcare, guided by user feedback and outcome monitoring. These hubs exemplify how emotional outcomes can be formalised through intersectoral governance and public–private partnerships[39].

The UK’s Lifetime Neighbourhoods strategy incorporates co-design principles into local planning. It establishes resident panels and supports participatory evaluation. These frameworks help align aging-related services with user needs through structured and inclusive procedures, rather than relying solely on top-down planning[40].

These models converge on three strategic lessons for China:

Institutional Design-Literacy: Design-literate governance requires policy actors to possess tools and competencies for co-creation. This should be formalised through professional training, cross-sectoral collaboration, and evaluative rubrics[41].

Emotional Infrastructure: Emotional care should be governed through dedicated instruments—such as wellbeing budgets, community counselling schemes, and metrics tracking loneliness or trust[42][43].

Place-Sensitive Engagement: Procedural flexibility and decentralised responsibility, as in Japan’s model, are vital for accommodating spatial diversity while maintaining care consistency.

To move beyond its infrastructure-centric paradigm, China must reconfigure its instrument mix by embedding these demand-side tools into legal mandates, budgetary structures, and procurement frameworks. Tools supporting emotional and participatory outcomes must no longer remain peripheral—they should become integral to the operational grammar of aging governance.

### 4.4 Towards Design-Literate, Health-Responsive aging Policy

Reframing China’s aging policy through a design-literate lens entails bridging spatial, emotional, and procedural dimensions. This transformation requires a paradigmatic shift from infrastructural output to dignity-centred service delivery.

Public sector capacity for service design should be progressively institutionalised. Experiences from Scandinavian and UK contexts suggest that embedding design units within government structures can foster iterative innovation and facilitate co-production with service users.

Emotional infrastructure could be addressed through dedicated policy instruments, including community-based mental health networks, longitudinal wellbeing monitoring, and targeted emotional-care funds. These measures are particularly relevant for tackling social isolation, psychological stress, and digital marginalisation among older populations. Participatory governance may be more effectively codified as a substantive policy function, rather than remaining at the level of symbolic consultation. Mechanisms such as citizen assemblies, feedback platforms, and policy pilot zones offer practical means to integrate user voices into decision-making and service design.

China’s experience in scaling physical upgrades—such as age-friendly renovations and smart infrastructure—provides a solid foundation. The challenge now lies in translating this capacity into holistic governance structures that support aging in place, intergenerational cohesion, and spatial justice. This shift would not only strengthen domestic aging services but also offer transferable frameworks for other Global South regions facing similar demographic pressures.

## 5. Conclusion

### 5.1 Reframing aging Governance through Design-Literate Instruments

This study employed a two-dimensional analytical framework that integrates policy instruments with age-friendly service design dimensions to examine structural misalignments within China’s community-level aging governance. Drawing on qualitative content analysis of 48 national-level policy documents, word frequency mapping, co-occurrence matrices, and empirical insights from fieldwork conducted in Zhenzhu Community, the analysis identified four interrelated governance deficits: A structural overreliance on supply-oriented instruments; Limited operational articulation of demand-driven and emotionally responsive service functions; Weak integration between spatial planning mechanisms and health-enabling governance functions; The absence of institutionalised coordination mechanisms across administrative domains.

The findings indicate that China’s aging policy system remains predominantly grounded in a material infrastructure paradigm. While the scale and frequency of physical improvements have increased, affective, participatory, and health-responsive dimensions continue to be institutionally marginalised. This reflects a systemic misalignment between normative commitments—such as dignity, inclusion, and emotional well-being—and the operational tools and routines deployed in policy implementation.

Empirical evidence from Zhenzhu Community illustrates this instrumental disjunction. Despite visible investment in physical infrastructure—such as accessible walkways and multifunctional service hubs—emotional services remain fragmented and episodic, with limited responsiveness to users’ lived experiences. The lack of formal feedback mechanisms further constrains adaptive policy revision. These deficiencies underscore the necessity of equipping aging policy with instruments capable of addressing the emotional, relational, and participatory dimensions of later life, rather than privileging physical interventions alone.

### 5.2 Policy Implications: From Infrastructure Delivery to Relational Governance

To address these structural gaps and reorient aging governance towards inclusive, design-literate, and health-promoting systems, four strategic policy directions are proposed:

#### 5.2.1 Recalibrate the Instrument Mix through Demand-Oriented Tools

The current policy toolkit remains overly reliant on directive and supply-driven measures. A shift is required toward instruments that actively engage users in service co-production and adaptive planning. This includes:

Participatory design protocols;

Behavioural insight techniques (nudges); Community scenario planning;

User-generated service feedback loops.

These tools facilitate the identification of differentiated needs and ensure that policy implementation remains sensitive to aging heterogeneity across spatial, cultural, and institutional contexts.

#### 5.2.2 Institutionalise Cross-Sectoral and Multi-Level Coordination

aging governance must transcend administrative silos through formalised coordination platforms. Key mechanisms include:

Joint policy taskforces linking health, planning, social services, and housing;

Shared performance metrics that incentivise inter-agency collaboration;

Cross-sectoral budgeting models with flexible allocation mandates.

Such arrangements enable horizontal and vertical coherence, reduce duplication, and embed aging-responsive logic into broader urban and social policy frameworks.

#### 5.2.3 Embed Relational Service Criteria into Urban Renewal and Community Planning

Age-friendly service design dimensions should be codified within planning regulations and procurement criteria. Physical regeneration projects must be assessed not only through spatial standards but also through relational indicators, such as:

Emotional well-being metrics;

Perceived accessibility and safety;

Intergenerational interaction indices.

This requires interdisciplinary evaluation frameworks that incorporate social care professionals, designers, planners, and community stakeholders from the project’s inception to implementation.

#### 5.2.4 Operationalise Intangible Values through Enforceable Design Standards

Normative principles—such as dignity, empathy, and inclusion—must be translated into regulatory clauses, modular toolkits, and institutional mandates. This involves:

National design guidelines for age-responsive policymaking;

Procurement templates that reward co-creation and emotional impact;

Legal instruments that mandate citizen engagement in service reform,

Through this approach, emotional infrastructure and participatory governance cease to be aspirational concepts and become formal domains of governance and accountability.

In sum, aging policy should evolve from an infrastructure-centric paradigm toward a design-literate model of governance that is relationally embedded, emotionally responsive, and procedurally adaptive. Built environments should not merely serve functional needs but act as platforms for psychosocial resilience, intergenerational solidarity, and health equity. China’s leadership in infrastructure investment provides a foundation for such a transition, but future policy success will depend on recalibrating instrument deployment toward dignity-centred and participatory governance. This lesson extends beyond the Chinese context and is of particular relevance to countries in the Global South confronting similar demographic shifts under constrained institutional capacities.

## Data Availability

All relevant data are within the manuscript and its Supporting Information files.

## Acknowledgments

I would like to express my sincere gratitude to Associate Professor Chen Sijing of Anhui University for her invaluable insights during the conceptualisation of this study, as well as her guidance in calibrating the independent double coding and her thorough review of the manuscript. I would also like to thank the Zhenzhu Community in Feixi County, Hefei City, Anhui Province, for their support and participation in the fieldwork.

## Ethics Statement

This study draws primarily on publicly accessible national-level policy documents and qualitative textual analysis. The fieldwork component, conducted in Zhenzhu Community, involved non-intrusive site observations and informal conversations with community staff, centring exclusively on institutional arrangements and policy implementation practices. No personally identifiable information or sensitive health-related data were collected at any stage. All engagements were undertaken with verbal informed consent, and participants were explicitly informed of the academic and non-commercial purpose of the study.

In line with institutional protocols and international ethical standards for minimal-risk social research, formal ethics review and approval were deemed unnecessary.

It should be noted that the study’s analytical scope is constrained by its reliance on central policy texts and a single case site. Future research may benefit from incorporating comparative local cases and longitudinal health-outcome data to enhance generalisability and empirical depth.

## Declaration of generative AI and AI-assisted technologies in the writing process

During the preparation of this work, I used ChatGPT in order to improve the language clarity and readability. After using this tool, I reviewed and edited the content as needed and take full responsibility for the content of the publication.

## Supporting Information Captions

**S1 Appendix. Catalogue of National Policy Documents on Ageing (1993–2025)**

This appendix presents a comprehensive catalogue of 48 national-level policy documents on ageing issued between 1993 and 2025. Each entry includes the policy title, issuing agency, publication date, and official document number, serving as the primary data source for NVivo coding and content analysis.

**S2 Appendix. NVivo Coding Structure Template.**

This appendix provides the complete coding structure used in the NVivo analysis, detailing node classifications for both policy instruments and service design dimensions.

**S3 Appendix. Community Observation Record Summary Table**

This appendix summarises field observation data collected in Zhenzhu Community, Feixi County, during 2024. It records key observations on spatial layout, service accessibility, interdepartmental coordination, older residents’ engagement, and emotional support mechanisms. These records complement the policy document analysis by offering empirical insight into the practical implementation of ageing-related services at the community level.

**S4 Appendix. Table of Source Data for Heatmap Visualization**

This appendix provides the underlying matrix data used for constructing the co-occurrence heatmap of policy instruments and service design dimensions. The table includes frequency counts derived from NVivo matrix coding across 48 national-level ageing policy documents (1993–2025), highlighting how each of the 22 policy instruments intersect with six design dimensions. These raw data form the empirical foundation for the visual analysis presented in Figure 2 of the main text.

